# Genetic influences for distinct impulsivity domains are differentially associated with early substance use initiation: Results from the ABCD Study

**DOI:** 10.1101/2025.04.14.25325687

**Authors:** Ethan Kinstler, Aaron J. Gorelik, Sarah E. Paul, Adamya Aggarwal, Emma C. Johnson, Melissa A. Cyders, Arpana Agrawal, Ryan Bogdan, Alex P. Miller

## Abstract

**Background:** Impulsivity is among the strongest correlates of substance involvement and its distinct domains (e.g., sensation seeking, urgency) are differentially correlated, phenotypically and genetically, with unique substance involvement stages. Understanding whether polygenic influences for distinct impulsivity domains are differentially predictive of early substance use initiation, a major risk factor for later problematic use, will improve our understanding of the role of impulsivity in addiction etiology.

**Methods:** Data collected from participants (*n*=4,808) of genetically-inferred European ancestry enrolled in the Adolescent Brain Cognitive Development Study^SM^ were used to estimate associations between polygenic scores (PGS) for UPPS-P impulsivity domains (i.e., sensation seeking, lack of premeditation/perseverance, and negative/positive urgency) and substance (i.e., any, alcohol, nicotine, cannabis) use initiation before age 15. Mediation models examined whether child impulsivity (ages 9-11) mediated links between PGSs and substance use initiation.

**Results:** Sensation seeking PGS was significantly associated with any substance and alcohol use initiation (ORs>1.10, *p*s_FDR_<0.006). Lack of perseverance and urgency (negative/positive) PGSs were nominally associated with alcohol and nicotine use initiation, respectively (ORs>1.06, *p*s<0.05, ps_FDR_>0.05). No significant associations were observed for lack of premeditation PGS or cannabis use initiation. Measured impulsivity domains accounted for 5-9% of associations between UPPS-P PGSs and substance use initiation.

**Conclusions:** Genetic influences for distinct impulsivity domains have differential associations with early substance use initiation with sensation seeking showing the most robust associations. Evaluating genetic influences for distinct impulsivity domains can yield valuable etiologic insight into the earliest stages of substance involvement that may be missed when adopting broad impulsivity definitions.

## Introduction

Impulsivity reflects a broad tendency to act quickly, without consideration of other options or consequences, in response to temporally salient external stimuli or internal drives (Whiteside & Lynam, 2001). It is among the strongest correlates of substance involvement (e.g., initiation of substance use, escalating use, and problematic use; de Wit, 2009), and is conceptualized as a predispositional risk factor for substance use (Verdejo-García et al., 2008). Consistent with this notion, impulsivity is moderately heritable and genetically correlated with substance involvement (e.g., use, problematic use; Sanchez-Roige et al., 2023), and elevated impulsivity during childhood and adolescence predicts substance use initiation and escalating use (McGue et al., 2001; Reynolds et al., 2003). Understanding how genetic influences for impulsivity are associated with early substance use initiation during the critical transition to adolescence may advance etiologic theories of addiction.

Contemporary conceptualizations view “impulsivity” as an amalgamation of partially overlapping, but distinct constructs, including sensation seeking (i.e., tendency to seek novel experiences), lack of perseverance (i.e., tendency to not finish what is started), lack of premeditation (i.e., tendency to act without forethought), and negative and positive urgency (i.e., tendency for rash action during elevated negative or positive mood states, respectively), rather than as a unitary construct (Cyders & Smith, 2008; Whiteside & Lynam, 2001). As studies have consistently shown that distinct impulsivity domains are differentially associated with substance use outcomes, separate consideration of these traits is necessary to further our understanding of the role each plays in the development of substance use (Strickland & Johnson, 2021). For example, sensation seeking is more strongly tied to both substance use initiation and use phenotypes (i.e., consumption, frequency) than other domains of impulsivity (Peeters et al., 2014; Stamates & Lau-Barraco, 2017; Stautz & Cooper, 2013). By contrast, urgency, both positive and negative, and lack of premeditation have been more strongly linked to problematic use (e.g., prolonged and disordered use) and unfavorable treatment outcomes compared to other domains (Coskunpinar et al., 2013; Cyders & Smith, 2008; McCarty et al., 2017; Stamates & Lau-Barraco, 2017; Verdejo-García et al., 2007). Different associations between impulsivity domains and substance involvement suggest that sensation seeking may be more etiologically relevant in earlier stages of substance use, whereas urgency and lack of premeditation may be more relevant in the transition to substance use disorders (SUDs). Understanding these associations can refine developmentally-appropriate prevention and intervention strategies (Halcomb et al., 2019; Kozak et al., 2019; Tomko et al., 2016).

All impulsivity domains are moderately, though variably, heritable (*h*^2^=0.27-0.50) and have unique and shared genetic components (Bezdjian et al., 2011; Friedman et al., 2020; Sanchez-Roige et al., 2023). Indeed, genome-wide association studies (GWAS) have revealed that while some impulsivity domains are highly genetically correlated (e.g., negative and positive urgency [rG=0.78]), others are moderately (e.g., lack of perseverance and premeditation [rG=0.50]), or even weakly correlated (e.g., sensation seeking and lack of perseverance [rG=0.01]; Sanchez-Roige et al., 2023). Variability in genetic correlations and multi-omic sources of overlap across impulsivity domains are also found with substance use and SUDs (Miller & Gizer, 2024; Sanchez-Roige et al., 2023; Vilar-Ribó et al., 2025). Specifically, while lack of premeditation and negative and positive urgency display similar genetic correlations with substance use and SUDs (rG=0.38-0.46), sensation seeking is more strongly genetically correlated with substance use (rG=0.27) *vs*. SUDs (rG=0.10; Miller & Gizer, 2024; Vilar-Ribó et al., 2025). Additionally, sensation seeking polygenic scores (PGSs), which provide a cumulative measure of GWAS-based genetic propensity (Wray et al., 2021), have been associated with early binge drinking frequency across independent samples over and above alcohol-related and other impulsivity domain PGSs (Ksinan et al., 2019; Lannoy et al., 2023; Miller, Spychala, et al., 2024). Further, evidence suggests sensation seeking may be a plausible mechanism through which genetic influences on substance involvement emerge as sensation seeking partially mediates links between polygenic scores for risk taking and alcohol involvement to alcohol use behavior among developmental samples (Ksinan et al., 2019; Lannoy et al., 2023). These mediational models suggest that a portion of genetic influences on drinking patterns in development may be attributable to direct expression of distinct impulsivity domains.

In the present study we examined the extent to which genetic influences for distinct impulsivity domains (i.e., sensation seeking, lack of perseverance, lack of premeditation, and negative & positive urgency) are associated with substance use initiation (i.e., any, alcohol, nicotine, cannabis) at an early age (i.e., <15 years) in 4,808 children in the Adolescent Brain Cognitive Development Study^SM^ (ABCD Study®) who most closely resemble European ancestry populations from genetic reference panels. We hypothesized that elevated genetic influences for sensation seeking would be the strongest predictor of early substance use initiation but that genetic influences for other domains would also be positively associated with early substance use initiation. We additionally tested whether variability in expressed impulsivity across domains may plausibly account for associations between genetic influences for impulsivity domains and substance use initiation. We hypothesized that the expression of domain-specific impulsivity would account for an appreciable portion of the association between genetic influences for each domain and substance use initiation, suggesting effects of domain-specific genetic influences for impulsivity on early substance use initiation partially operate through corresponding phenotypic expression, with smaller contributions arising from the expression of other domains.

## Methods

### Participants

The Adolescent Brain Cognitive Development Study^SM^ (ABCD Study®) is a longitudinal study of child and adolescent brain and behavioral development that recruited 11,875 9-11 year-old children (born 2005-2009) at baseline in 2016-2018 from 21 U.S. research sites (Volkow et al., 2018). It includes a family-based component, in which some participants were twins, triplets, and non-twin siblings. Parents/caregivers provided written informed consent, and children assent, to a research protocol approved by the institutional review board at each site (https://abcdstudy.org/sites/abcd-sites.html). Phenotypic data (*n*_analytic_=4,808) from data release 5.1 were obtained from the National Institute of Mental Health Data Archive (NDA, https://nda.nih.gov/abcd). Analyses were limited to individuals with non-missing impulsivity and substance use data whose genetic ancestry was similar to European ancestry reference populations due to the lack of relevant discovery GWAS in other ancestries and evidence that genomic influences may differ across ancestries. Applying polygenic scores to samples that are not similar to the ancestry of the original GWAS may contribute to ancestral health disparities by producing false positive and negative results (Martin et al., 2017, 2019).

### Impulsivity Domains

Impulsivity domains were measured using the 20-item abbreviated UPPS-P Impulsive Behavior Scale-Youth Version, which provides valid and reliable assessment of these traits in youth (Cyders et al., 2007; Watts et al., 2020). The UPPS-P captures the following five impulsivity domains, each with four items: (1) sensation seeking, (2) lack of perseverance, (3) lack of premeditation, (4) negative urgency, and (5) positive urgency (**Table 1**). Items are rated on a Likert scale of 1 (agree strongly) to 4 (disagree strongly); scores are reverse-coded, as needed, and summed, so that higher scale scores indicate more impulsivity.

**Table 1.** Sample descriptives by substance use initiation group.

| | Substance Naïve<br>($n=2,910$) | Substance Use Initiation<br>($n=1,898$) |
| --- | --- | --- |
| **Demographics** |  |  |
| Baseline age (years)(mean±SD) | 9.89±0.63 | 9.99±0.63 |
| Female sex (n)(%) | 1,421 (48.8%) | 835 (44.0%) |
| **Baseline impulsivity traits (mean±SD)** |  |  |
| Sensation seeking | 9.67±2.59 | 10.40±2.60 |
| Lack of perseverance | 6.94±2.17 | 7.26±2.24 |
| Lack of premeditation | 7.68±2.24 | 8.23±2.36 |
| Negative urgency | 8.23±2.54 | 8.74±2.53 |
| Positive urgency | 7.54±2.75 | 7.90±2.82 |
| **Impulsivity PGS (% in top decile [95% CI])** |  |  |
| SS-PGS | 9.00 [8.02, 10.10] | 11.50 [10.2, 13.10] |
| LPERSEV-PGS | 9.93 [8.90, 11.10] | 10.10 [8.84, 11.60] |
| LPREMED-PGS | 9.73 [8.70, 10.90] | 10.40 [9.13, 11.90] |
| NU-PGS | 9.62 [8.60, 10.70] | 10.60 [9.28, 12.10] |
| PU-PGS | 9.18 [8.18, 10.30] | 11.30 [9.93, 12.80] |

*Note:* PGS=polygenic score; SS=sensation seeking; LPERSEV=lack of perseverance; LPREMED=lack of premeditation; NU=negative urgency; PU=positive urgency.

### Substance Use Initiation

Annual in-person (i.e., baseline and follow-ups at 1-year [FU1], 2-year [FU2], and 3-year [FU3]) and mid-year phone (i.e., 6-month [FU0.5], 18-month [FU1.5], 30-month [FU2.5]) interviews assessed substance use (Lisdahl et al., 2018). Youth who endorsed use at any assessment from baseline to FU3 (i.e., lifetime use) were included in substance use initiation groups (Miller, Baranger, et al., 2024). Youth reporting substance use only in the context of religious ceremonies were coded as having missing data for substance use to restrict comparisons to use outside these settings (*n*_alcohol religious ceremonies_=432; *n*_nicotine religious ceremonies_=18). See **Supplementary Table 1** for a list of study release variables used to code substance use initiation.

### Alcohol use initiation

(*n*=1,778) was defined as ‘sipping’ (*n*=1,658 with no full drink) or ‘full drinks’ (*n*=120) of alcohol. This definition of any use, inclusive of sipping, was used (1) to provide more consistent definitions of use across substances, (2) due to prior evidence that ‘sipping’ is similarly associated with externalizing as full drinks in youth (Watts et al., 2021, 2024), (3) due to power concerns with modeling initiation at different levels given low endorsements, and (4) to retain consistency with other ABCD study definitions (Miller, Baranger, et al., 2024; Watts et al., 2021, 2024). ***Nicotine use initiation*** (*n*=201) was defined as use of nicotine products in any form. ***Cannabis use initiation*** (*n*=75) was defined as use of cannabis in any form, except synthetic cannabis or cannabis-infused alcoholic drinks, which were included under any substance use. ***Any substance use initiation*** (*n*=1,898) was defined as alcohol, nicotine, or cannabis use initiation, or undirected/recreational use of any other substances (*n*=87: *n*_inhalants_=27, *n*_prescription sedatives_=17, *n*_stimulants_=16, *n*_synthetic cannabis_ =13, *n*_OTC cough/cold medicine_=7, *n*_opioids_=7, *n*_hallucinogens_=4, *n*_other_=6; **Table 1**). ***Substance naïve youth*** (*n*=2,910) endorsed no substance use from baseline to FU3 and had non-missing FU3 data to protect against misclassification of participants with an unknown FU3 status (*n*=327). Notably, there was considerable overlap among initiation of alcohol, nicotine, and cannabis in our sample with alcohol use initiation representing the vast majority of substance use initiation (94%; **Supplementary Figure 1**).

### Polygenic Scores

Saliva samples were genotyped on the Affymetrix Smokescreen array (Baurley et al., 2016) by the Rutgers University Cell and DNA Repository (company now known as “Sampled”; https://sampled.com/). Genotyped calls were aligned to GRC37 (hg19). The Rapid Imputation and COmputational PIpeLIne for Genome-Wide Association Studies (RICOPILI; (Lam et al., 2019) was used to perform quality control on the 11,099 individuals with available ABCD Study phase 3.0 genotypic data, using default parameters. The 10,585 individuals who passed QC checks were subjected to principal component analysis (PCA) in RICOPILI to map the genomic ancestry of these individuals to the 1000 Genomes reference panel, resulting in a PCA-selected subset of 5,556 individuals who most resembled the 1000 Genomes European reference population. Only individuals of genetically-inferred European ancestry were analyzed (see **Participants**). The Trans-Omics for Precision Medicine (TOPMed) imputation reference panel was used for genotype imputation (Taliun et al., 2021). Imputation dosages were converted to best-guess hard-called genotypes; only single nucleotide polymorphisms (SNPs) with Rsq > 0.8 and MAF > 0.01 were kept for PGS computation.

GWAS summary statistics, drawn from the largest discovery UPPS-P GWAS available (Sanchez-Roige et al., 2023), were used to develop PGS for each of the five UPPS-P domains (h^2^_SNP_=0.06-0.10; N=132,132-133,517): sensation seeking (SS-PGS), lack of perseverance (LPERSEV-PGS), lack of premeditation (LPREMED-PGS), negative urgency (NU-PGS), and positive urgency (PU-PGS). Participants in this GWAS were all research-consented adults (median age=54) of European ancestry from the consumer genetics and research company 23andMe, Inc. and responded to a research survey including the 20-item short-form version of the UPPS-P (Cyders et al., 2014; Sanchez-Roige et al., 2023). SNP weights for PGSs were generated using PRS-CS (v1.0.0), a Bayesian polygenic prediction method that infers posterior SNP effects sizes under continuous shrinkage priors using GWAS summary statistics and an external linkage disequilibrium reference panel (1000 Genomes Project phase 3; Ge et al., 2019). The ‘auto’ feature of PRS-CS was used to learn the global shrinkage parameter from the data using a fully Bayesian approach with 10,000 Markov chain Monte Carlo (MCMC) iterations, a burn-in sample of 5,000, and a thinning interval of 5. The --score function in PLINK (v2.0; Chang et al., 2015) was used to compute PGSs in the ABCD sample using PRS-CS-derived weights for 866,844 overlapping HapMap3 SNPs with the 1000 Genomes European ancestry sample used as the reference panel for linkage disequilibrium.

### Statistical Analysis

All continuous variables were z-scored prior to analyses. Substance use initiation groups (i.e., any [*n*=1,898], alcohol [*n*=1,778], nicotine [*n*=187], cannabis [*n*=75] *vs*. substance naïve [*n*=2,910]) were logistically regressed on the five UPPS-P PGSs (i.e., SS-PGS, LPERSEV-PGS, LPREMED-PGS, NU-PGS, PU-PGS) using separate models (i.e., for each UPPS-P PGS and substance use initiation group). All analyses were conducted using maximum likelihood with robust standard errors (MLR) estimation in Mplus 8.11 (Muthén & Muthén, 1998-2024) via the MplusAutomation R package (v1.1.1; Hallquist & Wiley, 2018). Fixed effect covariates included: baseline age, age-squared, sex, familial relationship (i.e., sibling, twin, triplet), and 10 genomic principal components (PCs). Data were nested within recruitment site (i.e., stratum) and families (i.e., cluster) to account for the non-independence of data and adjust standard errors. Pubertal status as an additional fixed effect covariate was examined *post hoc* and did not meaningfully impact observed associations (**Supplementary Table 2**). Multiple testing was adjusted using a 5% false discovery rate (FDR) correction separately within each substance use initiation contrast (i.e., any *vs*. substance naïve, alcohol *vs*. substance naïve, nicotine *vs*. substance naïve, cannabis *vs*. substance naïve) for the five UPPS-P PGSs tested.

For any nominally significant association observed between UPPS-P PGSs and substance use initiation variables, we conducted two additional analyses. *First*, we conducted multiple regression with all UPPS-P PGSs included simultaneously to evaluate whether associations were independent of other UPPS-P PGSs. *Second*, we examined the degree to which UPPS-P self-reports measured at baseline accounted for associations between UPPS-P PGSs and substance use initiation using parallel mediation models. Here, in each model, baseline UPPS-P scale scores for each domain were entered as parallel mediators for associations between UPPS-P PGSs and substance use initiation. Monte Carlo integration was used to account for missing baseline UPPS-P measures (*n*=4). Significance of indirect associations (i.e., mediation) were assessed using 95% confidence intervals computed via Monte Carlo simulation of estimated model parameters and their associated asymptotic sampling covariance matrices (20,000 simulations) using the *monteCarloCI* function in the semTools R package (v0.5-6; Jorgensen et al., 2022; MacKinnon et al., 2004; Preacher & Selig, 2012). Estimates of the proportion of total effects (i.e., sum of direct and indirect effects of PGSs) on substance use initiation significantly mediated by baseline UPPS-P measures were obtained by calculating the ratio of indirect effect estimates for each significant path to the total effect estimate.

## Results

### Any Substance Use initiation

Higher SS-PGS was associated with a greater likelihood of any substance use initiation (*n*_*any*_*=*1,898; OR=1.108, 95% CI [1.042, 1.179], *p*=0.001, *p*_FDR_=0.005; **Figure 1; Supplementary Table 3**), which remained significant when including all UPPS-P PGS simultaneously in the model (OR=1.106, 95% CI [1.035, 1.182], *p*=0.003, *p*_FDR_=0.015; **Supplementary Table 4**). The parallel mediation model revealed that elevated baseline sensation seeking (β_indirect_=0.009, 95% CI [0.002, 0.017]) and lack of premeditation (β_indirect_=0.006, 95% CI [0.001, 0.011]) indirectly linked heightened SS-PGS to any substance use initiation accounting for 8.9% and 5.3% of this association, respectively (**Figure 2A; Supplementary Table 5**).

**Figure 1.**
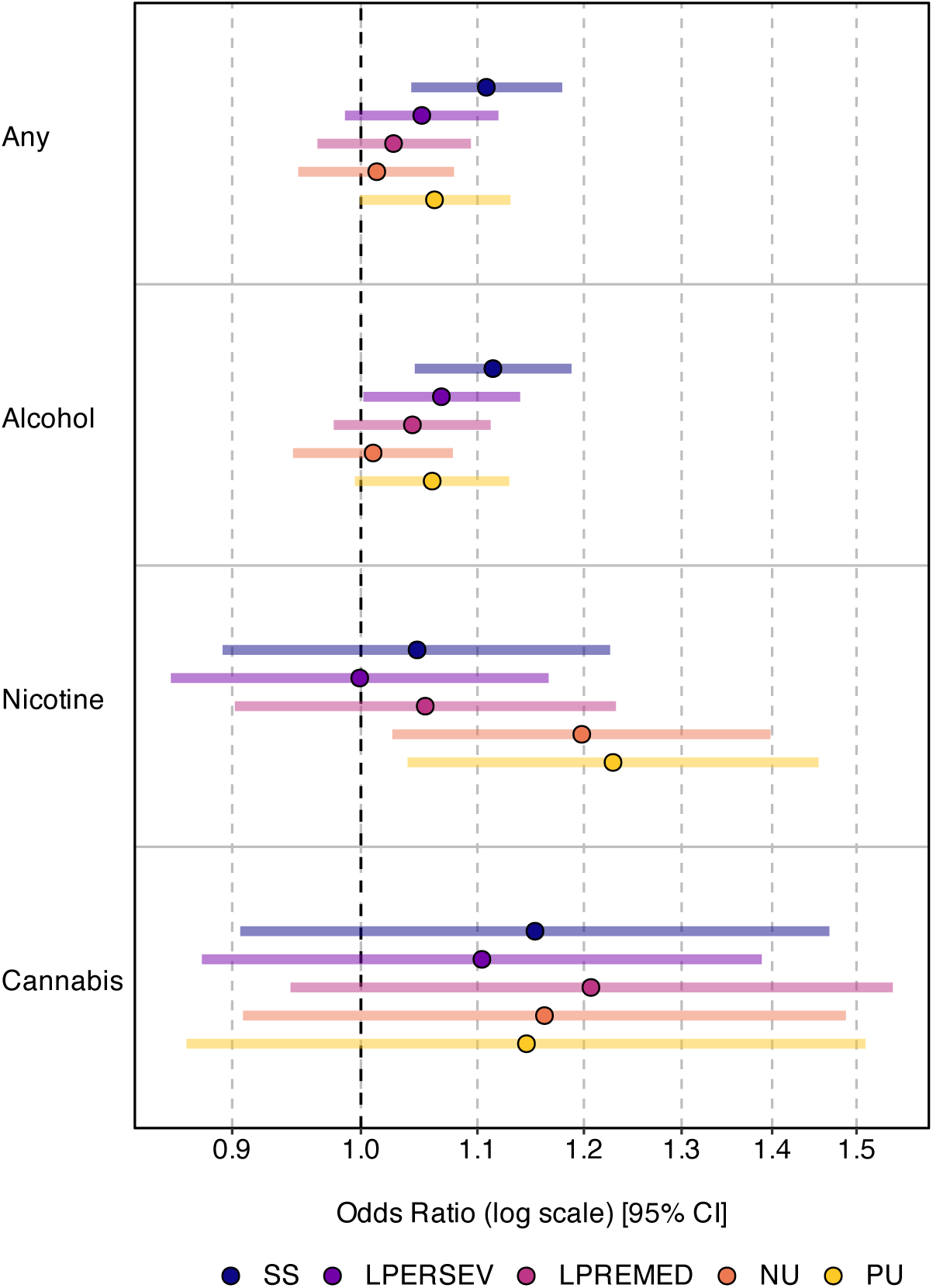
Associations between UPPS-P PGSs and substance use initiation. Odds ratios and 95% confidence intervals, presented on a log scale, for associations between each UPPS-P PGS and substance use variable. SS=sensation seeking; LPERSEV=lack of perseverance; LPREMED=lack of premeditation; NU=negative urgency; PU=positive urgency.

**Figure 2.**
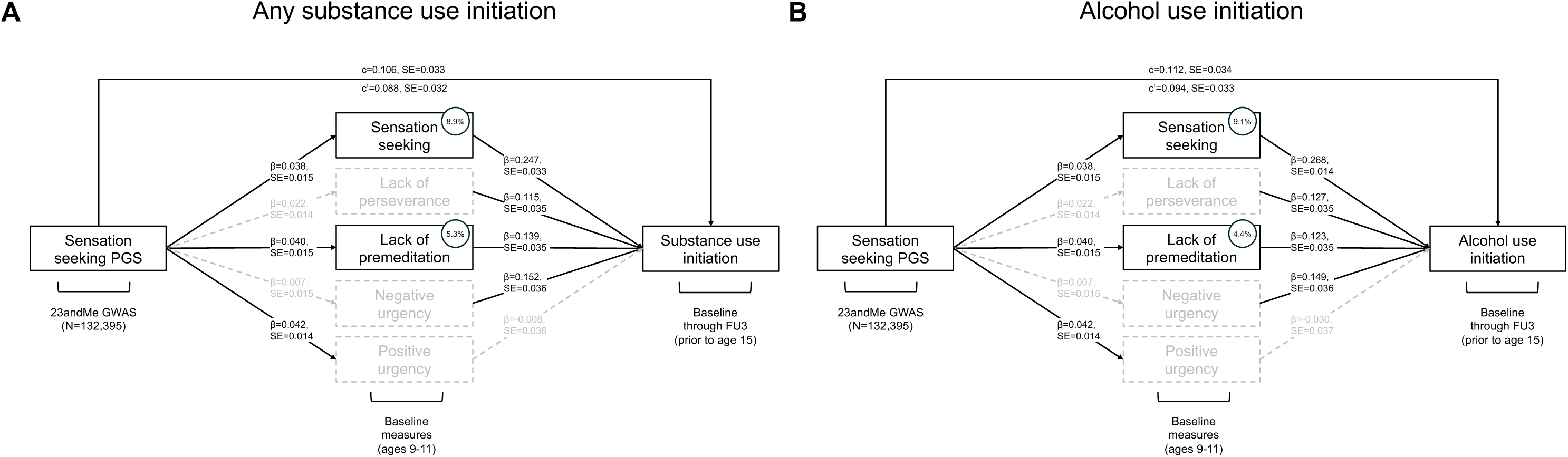
Sensation seeking and lack of premeditation indirectly link sensation seeking PGS to (A) any substance use and (B) alcohol use initiation. Mediational models including all measured UPPS-P domains in parallel. Standardized regression coefficients (β) and standard errors (SE) are reported along each path. Grey dashed lines indicate individual paths and/or mediation effects with 95% CIs overlapping with 0 (i.e., non-significant). Numbers in circles represent the proportion of the association between SS-PGS and the substance use initiation variable that is accounted for by the measured UPPS-P domain (i.e., sensation seeking and lack of premeditation accounted for 8.9 and 5.3% of association between SS-PGS and any substance use initiation, respectively, and 9.1% and 4.4% of the association with alcohol use initiation, respectively).

### Alcohol Use Initiation

Higher SS-PGS was significantly associated with a greater likelihood of alcohol use initiation (*n*_*alcohol*_*=*1,778, SS-PGSs: OR=1.114, 95% CI [1.045, 1.188], *p*=0.003, *p*_FDR_=0.005; **Figure 1**; **Supplementary Table 3**), which remained significant when including all UPPS-P PGS simultaneously in the model (OR=1.111, 95% CI [1.039, 1.189], *p*=0.002, *p*_FDR_=0.010; **Supplementary Table 4**). Baseline sensation seeking (β_indirect_=0.010, 95% CI [0.002, 0.019]) and lack of premeditation (β_indirect_=0.005, 95% CI [0.001, 0.010]) indirectly linked heightened SS-PGS to alcohol use initiation accounting for 9.1% and 4.4% of this association, respectively (**Figure 2B; Supplementary Table 6**). Higher LPERSEV-PGS was also associated with a greater likelihood of alcohol use initiation (OR=1.680, 95% CI [1.002, 1.139], *p*=.043, *p*_FDR_=.107, **Figure 1**); however, this association did not survive FDR correction. Baseline lack of perseverance (β_indirect_=0.005, 95% CI [0.001, 0.011]) and lack of premeditation (β_indirect_=0.004, 95% CI [0.001, 0.009]) indirectly linked heightened LPERSEV-PGS to alcohol use initiation accounting for 7.7% and 6.4% of these associations, respectively (**Supplementary Figure 2; Supplementary Table 7**).

### Nicotine Use Initiation

Higher NU-PGS and PU-PGS were associated with a greater likelihood of nicotine use initiation (*n*_*nicotine*_*=*187; NU-PGSs: OR=1.198, 95% CI [1.026, 1.398], *p=*0.022, *p*_FDR_=0.055; PU-PGSs: OR=1.229, 95% CI [1.039, 1.454], *p*=0.016, *p*_FDR_=0.055; **Figure 1**; **Supplementary Table 3**); however, these associations did not survive FDR correction. Neither NU-PGS nor PU-PGS were a significant independent predictor of nicotine use initiation when entered into the model simultaneously (NU-PGSs: OR=1.210, 95% CI [0.938, 1.339], *p=*0.209, *p*_FDR_=0.522; PU-PGSs: OR=1.163, 95% CI [0.953, 1.420], *p*=0.839, *p*_FDR_=0.994; **Supplementary Table 4**). Baseline negative urgency indirectly linked heightened NU-PGS to nicotine use initiation accounting for 5.3% of this association (β_indirect_=0.009, 95% CI [0.001, 0.020]; **Supplementary Figure 3; Supplementary Table 8**). Both baseline negative and positive urgency similarly indirectly linked heightened PU-PGS to nicotine use initiation (negative urgency-β_indirect_=0.010, 95% CI [0.002, 0.022]; positive urgency-β_indirect_=0.010, 95% CI [0.001, 0.022]) accounting for 5.1% and 5.2% of these associations, respectively (**Supplementary Figure 4; Supplementary Table 9**).

### Cannabis Use Initiation

No UPPS-P PGSs were significantly associated with cannabis use initiation (*n*_*cannabis*_=75; **Figure 1; Supplementary Table 3**).

## Discussion

Our study of genetic influences for impulsivity domains (i.e., sensations seeking, lack of perseverance, lack of premeditation, negative urgency, and positive urgency) and substance use initiation as children enter adolescence revealed three primary findings. *First*, consistent with evidence that sensation seeking is the impulsivity domain most strongly associated with substance use initiation and continued use in adolescence (Cappelli et al., 2020; Stautz & Cooper, 2013), SS-PGS was the most prominent polygenic predictor of substance use initiation. *Second*, NU-PGS and PU-PGS were both uniquely predictive of nicotine use initiation with largely overlapping effects. *Third*, phenotypically expressed domains of impulsivity accounted for 5-9% of the variance between PGSs and substance use initiation; these indirect effects were primarily characterized by domain congruency (e.g., sensation seeking mediating links between SS-PGS and substance use initiation), though incongruent domain mediation was also found (e.g., lack of premeditation also mediating the association between SS-PGS and substance use initiation). Collectively, our findings show that genetic influences for distinct impulsivity domains, as well as their expression, have differential associations with substance use initiation at an early age. More specifically, and consistent with prior phenotypic (Cappelli et al., 2020; Gunn & Smith, 2010; Jensen et al., 2017; Stautz & Cooper, 2013) and genetic (Miller, Spychala, et al., 2024; Vilar-Ribó et al., 2025) studies, these data further highlight sensation seeking in early substance use initiation. More broadly, these data emphasize the potential utility of GWAS of more precise dimensional phenotypes as well as the use of developmental samples in furthering our understanding of factors influencing early onset substance use.

### Sensation Seeking

Individuals with higher SS-PGS were more likely to have initiated any substance and/or alcohol use, and this effect was partially mediated by childhood phenotypic expression of sensation seeking and lack of premeditation, broadly consistent with prior studies demonstrating sensation seeking mediates PGS effects on alcohol use (Ksinan et al., 2019; Lannoy et al., 2023). This finding also aligns with prior phenotypic studies, which have found that sensation seeking most strongly predicts use at a young age, as well as initial and increasing substance (particularly alcohol) use in adolescence (Cappelli et al., 2020; Jensen et al., 2017; Stautz & Cooper, 2013). Additionally, as early initiation portends a greater risk for problematic use, it is plausible that at least some of the association between sensation seeking and later stages of substance involvement (e.g., problematic use, SUDs) may be attributable to its association with initiation (Doumas et al., 2019). Thus, sensation seeking may serve as an intermediate phenotype for problematic use through its association with earlier initiation, and our findings, which suggest that the earliest stages of this pathway may be influenced by genetic factors and related phenotypic expression, help clarify the mechanisms underlying this association. Contemporary conceptualizations of personality suggest that traits like sensation seeking exhibit substantial developmental change over the lifespan, especially from early childhood to adolescence (Khurana et al., 2018), and theoretical models postulate that these developmental personality changes are, in part, driven by genetically-programmed and environmentally-induced brain maturation (Casey et al., 2025; Harden et al., 2012). Thus, a more comprehensive picture of the genetic and neurobiological links between sensation seeking and early stages of substance involvement are beginning to emerge, in which sensation seeking is both phenotypically and genetically salient in substance use etiology.

### Urgency

NU-PGS and PU-PGS both exhibited nominally significant associations only with nicotine use initiation. While these findings did not survive FDR correction for multiple testing, existing literature highlights their potential relevance. Given links between negative and positive urgency and the development of alcohol and nicotine-related problems (Coskunpinar et al., 2013; Kale et al., 2018; Stamates & Lau-Barraco, 2017; Stautz & Cooper, 2013), the theorized role of negative urgency in withdrawal and craving processes in addiction (Zorrilla & Koob, 2019), and moderate genetic correlations with SUDs (Sanchez-Roige et al., 2023), these PGSs may reflect shared genetic variance with more severe stages of substance involvement. Consistent with this notion, fewer parents of ABCD Study participants report easy child access to nicotine relative to alcohol (Martz et al., 2022) and, in the current study, the majority (i.e., 73%) of participants endorsing nicotine use initiation also endorsed alcohol and/or cannabis use initiation, indicative of greater substance involvement. Lifetime nicotine use is also associated with elevated risk of dependence relative to lifetime use of other substances (Lopez-Quintero et al., 2011). As such, while speculative, the link between urgency PGSs and initiation of nicotine use by age 15 in this sample may reflect greater risk of progression through later stages of substance involvement. While urgency has been established as a risk factor for later nicotine dependence (Kale et al., 2018), the associations between genetic influences for negative/positive urgency and early nicotine use initiation in our study are novel, and more research in early experimentation is necessary.

These findings may also inform impulsivity domain conceptualizations. NU-PGS and PU-PGS associations with nicotine use initiation were overlapping, and the phenotypic expression of both urgency domains indirectly linked PU-PGS to nicotine use initiation. This is consistent with evidence that positive and negative urgency are highly genetically correlated (rG=.78; Sanchez-Roige et al., 2023) and may reflect a general urgency factor characterized by emotion-based rash action regardless of affective valence (Billieux et al., 2021). In contrast, more nuanced examinations of these genetic factors also suggest possible differential relations with substance use *vs*. SUDs (Vilar-Ribó et al., 2025). As current polygenic score modeling methods are not well-suited to leverage this level of nuance, it underscores the need for research that can capture unique genetic loadings across substance use development trajectories.

### Lack of Premeditation

Our finding that LPREMED-PGS was not associated with substance use initiation aligns with evidence that heightened lack of premeditation may be primarily associated later stages of substance involvement including heavy drinking and problematic alcohol use in emerging and young adulthood (Adams et al., 2012; Coskunpinar et al., 2013; Verdejo-García et al., 2007). As compared to sensation seeking, which exhibits stronger genetic associations with substance use *vs*. SUDs, lack of premeditation demonstrates equivalent genetic associations with both (Vilar-Ribó et al., 2025). Nevertheless, a portion of the association between SS-PGS and initiation of any substances or alcohol was mediated by baseline measures of lack of premeditation (~5%), suggesting some overlap between genetic influences on sensation seeking measured in adulthood and lack of premeditation in childhood. Such data are consistent with models highlighting cognitive control effects on both positive and negatively reinforcing aspects of substance involvement (Bogdan et al., 2023) as well as evidence that lack of premeditation potentiates the associations between sensation seeking and risky substance use (McCabe et al., 2015) and that sensation seeking is correlated with structural variability in regions critical to cognitive control (Holmes et al., 2016). However, as sensation seeking increases during late childhood and peaks during mid-adolescence but the development of premeditation protracts into later adolescence and young adulthood, it is possible that their phenotypic and genetic separation varies across development (Harden et al., 2012; Harden & Tucker-Drob, 2011; Steinberg, 2010). Further research is needed to examine the extent to which genetic influences for impulsivity domains may operate through divergent pathways across the lifespan.

### Limitations and Future Directions

Our study and results should be interpreted in the context of its limitations. *First*, highlighting calls for GWAS in more diverse ancestral populations (Corpas et al., 2025; Martin et al., 2019), GWAS of these traits in more diverse populations are needed to assess generalizability of findings and mitigate disparities. *Second*, consistent with epidemiological evidence that alcohol is typically the first substance used (Smit et al., 2018), the majority of substance use in our sample of children from the ABCD Study was alcohol use. Thus, whether our observed differential substance associations reflect true differences in substance-specific etiology and/or result from differential power (e.g., null associations between SS-PGS and nicotine and cannabis use initiation) is unknown. Relatedly, as the ABCD sample ages it will be important to model genetic influences of impulsivity domains on other substance involvement stages (e.g., escalating use, problematic use, relapse; (Miller, Spychala, et al., 2024; Paul et al., 2024). *Third*, consistent with polygenic scores for other complex behavioral phenotypes, our impulsivity domain PGSs were associated with behavioral outcomes (i.e., substance use initiation, reported impulsivity) with small effect sizes, suggesting that, while they can help inform phenotypic and genetic etiology, they have limited clinical utility at present (Bogdan et al., 2018; Ma & Zhou, 2021; Visscher et al., 2017). *Fourth*, some impulsivity traits, including sensation seeking, peak in post-pubertal adolescence before waning in adulthood (Harden & Tucker-Drob, 2011), and this pattern mirrors changes in substance use during this timeframe (Quinn & Harden, 2013). Impulsivity polygenic scores for the current study were derived using adult GWAS data (median age=54) that may not capture developmentally-specific genetic influences contributing to the differential expression of impulsivity across the lifespan (Bogdan et al., 2023). This notion is consistent with evidence that complex traits (e.g., alcohol consumption, asthma, BMI) may have distinguishable genetic influences across the lifespan (Couto Alves et al., 2019; Pividori et al., 2019; Thomas et al., 2024). *Finally*, environmental factors are robust correlates of substance involvement (McGue et al., 2000; Rhee et al., 2003) and some of the genetic effects we observed could emerge through gene-environment interplay. For example, associations may partially reflect passive gene-environment correlations, wherein parental genotype influences family environment (e.g., parental alcohol supply) impacting substance experimentation and later harmful drinking patterns (Chan et al., 2017; Gilligan et al., 2012). Research in well-powered samples that can carefully attend to methodological challenges associated with gene-environment influences are needed (Bogdan et al., 2018; Duncan & Keller, 2011).

## Conclusions

Impulsivity is one of the strongest correlates of substance involvement (de Wit, 2009). Phenotypically, decades of research have shown that distinct impulsivity domains are differentially associated with different stages of substance involvement (Coskunpinar et al., 2013; Stamates & Lau-Barraco, 2017; Stautz & Cooper, 2013). Emergent evidence suggests that domain-specific genetic influences for impulsivity demonstrate similar differential associations with substance use and SUDs (Sanchez-Roige et al., 2023; Vilar-Ribó et al., 2025). Consistent with this literature, our study demonstrates that genetic influences for distinct impulsivity domains are differentially associated with early substance use initiation in late childhood and early adolescence and that these effects are partially mediated by phenotypic expression of relevant impulsivity domains. In particular, genetic influences for sensation seeking emerged as robustly associated with substance use initiation. A greater understanding of the genetic influences for developmentally-dynamic traits, such as sensation seeking, and links to substance involvement provide crucial insights into the genetic etiology of early emergence of SUD risk factors, and how these disorders develop across the lifespan, potentially informing early intervention and prevention efforts.

## Data Availability

Data used in the preparation of this article were obtained from the Adolescent Brain Cognitive Development^SM^ (ABCD®) Study (https://abcdstudy.org), held in the NIMH Data Archive (NDA). This is a multisite, longitudinal study designed to recruit more than 10,000 children aged 9-10 and follow them over 10 years into early adulthood. The ABCD Study® is supported by the National Institutes of Health and additional federal partners under award numbers U01DA041048, U01DA050989, U01DA051016, U01DA041022, U01DA051018, U01DA051037, U01DA050987, U01DA041174, U01DA041106, U01DA041117, U01DA041028, U01DA041134, U01DA050988, U01DA051039, U01DA041156, U01DA041025, U01DA041120, U01DA051038, U01DA041148, U01DA041093, U01DA041089, U24DA041123, U24DA041147. A full list of supporters is available at https://abcdstudy.org/federal-partners.html. A listing of participating sites and a complete listing of the study investigators can be found at https://abcdstudy.org/consortium_members/. ABCD consortium investigators designed and implemented the study and/or provided data but did not necessarily participate in the analysis or writing of this report. This manuscript reflects the views of the authors and may not reflect the opinions or views of the NIH or ABCD consortium investigators. The ABCD data repository grows and changes over time. The ABCD data used in this report came from http://dx.doi.org/10.15154/1520591 (ABCD Annual Release 3.0) and http://dx.doi.org/10.15154/8873-zj65 (ABCD Annual Release 5.0). DOIs can be found at https://nda.nih.gov/abcd/abcd-annual-releases.html.

## Acknowledgements

Data for this study were provided by the Adolescent Brain Cognitive Development (ABCD) study, which was funded by award Nos. U01DA041022, U01DA041025, U01DA041028, U01DA041048, U01DA041089, U01DA041093, U01DA041106, U01DA041117, U01DA041120, U01DA041134, U01DA041148, U01DA041156, U01DA041174, U24DA041123, and U24DA041147 from the National Institutes of Health and additional federal partners (https://abcdstudy.org/federal-partners.html). The ABCD Study is supported by the National Institutes of Health and additional federal partners under award Nos. U01DA041048, U01DA050989, U01DA051016, U01DA041022, U01DA051018, U01DA051037, U01DA050987, U01DA041174, U01DA041106, U01DA041117, U01DA041028, U01DA041134, U01DA050988, U01DA051039, U01DA041156, U01DA041025, U01DA041120, U01DA051038, U01DA041148, U01DA041093, U01DA041089, U24DA041123, and U24DA041147. A full list of supporters is available at https://abcdstudy.org/federal-partners.html. We are thankful to ABCD Study participants and staff. This study made use of GWAS summary statistics data from 23andMe, Inc. (San Francisco, CA). We would like to thank the research participants and employees of 23andMe, Inc. for making this work possible.

## Financial Support

This study was supported by R01 DA054750 (RB & AA). Support was also provided by K01 AA031724 (APM), NSF DGE-213989 (AJG), F31 AA029934 (SEP), and K01 DA051759 (ECJ).

## Author Contributions

Mx. Kinstler and Dr. Miller had full access to all of the data in the study and take responsibility for the integrity of the data and the accuracy of the data analysis.

*Concept and Design:* Kinstler, Miller, Bogdan.

*Acquisition, analysis, or interpretation of data:* Kinstler, Miller, Bogdan.

*Drafting of the manuscript:* Kinstler, Miller, Bogdan.

*Critical review of the manuscript for important intellectual content:* Kinstler, Gorelik, Paul, Aggarwal, Johnson, Cyders, Agrawal, Bogdan, Miller.

*Statistical analysis:* Miller, Kinstler, Bogdan.

*Obtained funding:* Miller, Agrawal, Bogdan.

*Administrative, technical, or material support:* Paul, Gorelik, Agrawal, Johnson, Bogdan, Miller.

*Supervision:* Miller, Bogdan.

## Conflict of Interest Disclosures

None reported.

## Notes

### Competing Interest Statement

The authors have declared no competing interest.

### Author Declarations

Data used in the preparation of this article were obtained from the Adolescent Brain Cognitive Development^SM^ (ABCD®) Study (https://abcdstudy.org), held in the NIMH Data Archive (NDA). This is a multisite, longitudinal study designed to recruit more than 10,000 children aged 9-10 and follow them over 10 years into early adulthood. The ABCD Study is supported by the National Institutes of Health and additional federal partners under award numbers U01DA041048, U01DA050989, U01DA051016, U01DA041022, U01DA051018, U01DA051037, U01DA050987, U01DA041174, U01DA041106, U01DA041117, U01DA041028, U01DA041134, U01DA050988, U01DA051039, U01DA041156, U01DA041025, U01DA041120, U01DA051038, U01DA041148, U01DA041093, U01DA041089, U24DA041123, U24DA041147. A full list of supporters is available at https://abcdstudy.org/federal-partners.html. A listing of participating sites and a complete listing of the study investigators can be found at https://abcdstudy.org/consortium_members/. ABCD consortium investigators designed and implemented the study and/or provided data but did not necessarily participate in the analysis or writing of this report. This manuscript reflects the views of the authors and may not reflect the opinions or views of the NIH or ABCD consortium investigators. The ABCD data repository grows and changes over time. The ABCD data used in this report came from http://dx.doi.org/10.15154/1520591 (ABCD Annual Release 3.0) and http://dx.doi.org/10.15154/8873-zj65 (ABCD Annual Release 5.0). DOIs can be found at https://nda.nih.gov/abcd/abcd-annual-releases.html.

### Summary of Updates

First author name was misspelled.

