## Supplementary Figures for "Genetic influences for distinct impulsivity domains are differentially associated with early substance use initiation: Results from the ABCD Study"

**Table of Contents**

Supplementary Figure 1: Overlap among endorsement of alcohol, nicotine, and cannabis use initiation.
Supplementary Figure 2: Baseline UPPS-P mediating lack of perseverance PGS association with alcohol use initiation.
Supplementary Figure 3: Baseline UPPS-P mediating negative urgency PGS association with nicotine use initiation.
Supplementary Figure 4: Baseline UPPS-P mediating positive urgency PGS association with nicotine use initiation.

**Supplementary Figure 1. Overlap among endorsement of alcohol, nicotine, and cannabis use initiation.**

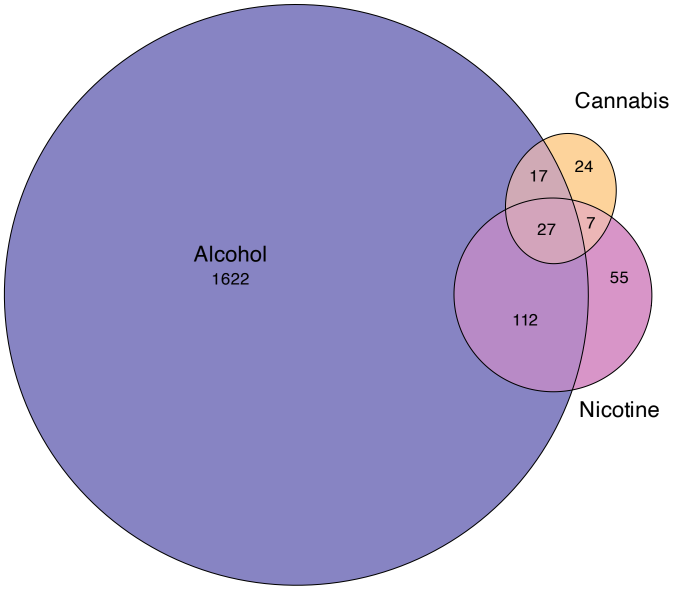


Overlap amongst participants endorsing alcohol (including both ‘sipping’ and ‘full drinks’), nicotine, and cannabis use initiation through follow-up year 3.

**Supplementary Figure 2. Baseline UPPS-P mediating lack of perseverance PGS association with alcohol use initiation.**

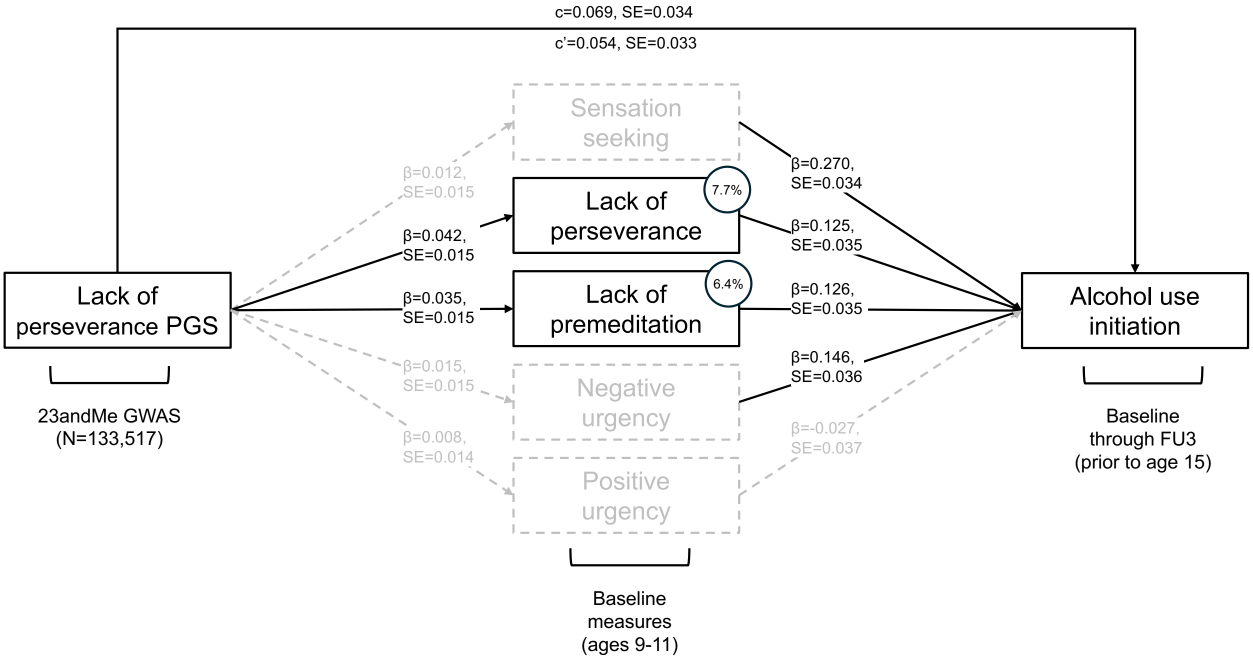


PGS = polygenic score; GWAS = genome-wide association study; c = total effect; c' = direct effect; FU = follow-up. Standardized regression coefficients (β) and standard errors (SE) are reported along each path. Grey dashed lines indicate individual paths and/or mediation effects with 95% CIs overlapping with 0 (i.e., non-significant). Estimates of proportion of the total effect mediated by baseline UPPS-P measures are included for all significant paths as numbers in circles (i.e., lack of perseverance and lack of premeditation accounted for 7.7 and 6.4% of association between lack of perseverance PGS and alcohol use initiation, respectively).

**Supplementary Figure 3. Baseline UPPS-P mediating negative urgency PGS association with nicotine use initiation.**

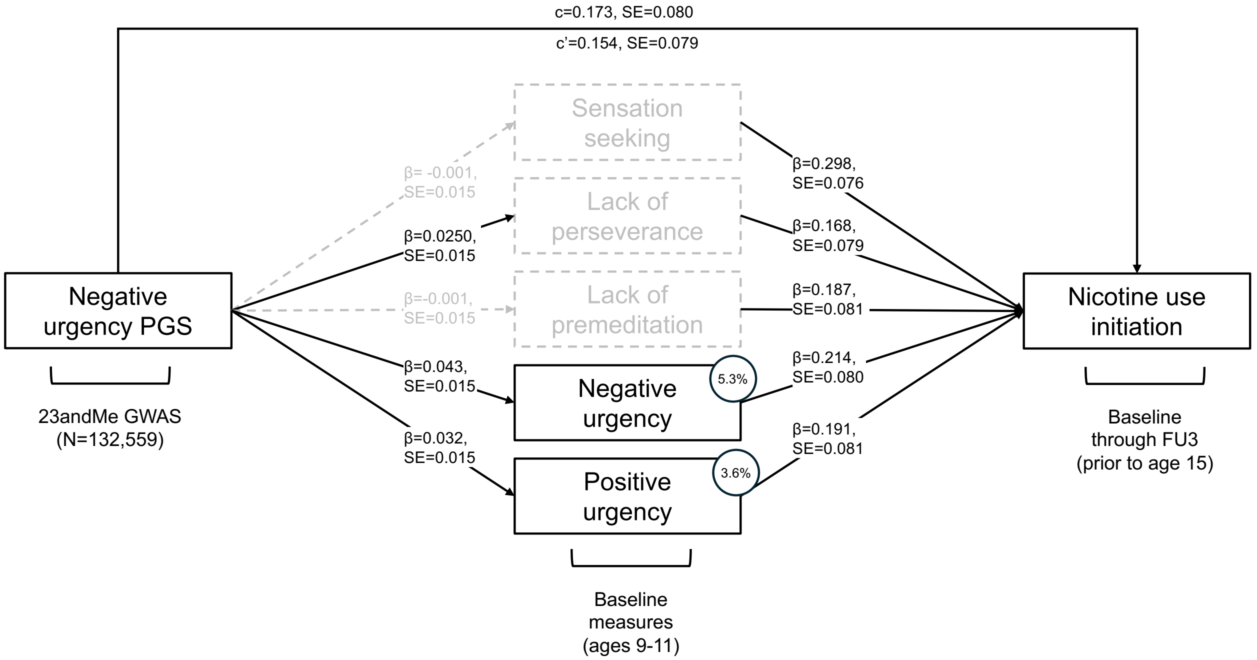


PGS = polygenic score; GWAS = genome-wide association study; c = total effect; c' = direct effect; FU = follow-up. Standardized regression coefficients (β) and standard errors (SE) are reported along each path. Grey dashed lines indicate individual paths and/or mediation effects with 95% CIs overlapping with 0 (i.e., non-significant). Estimates of proportion of the total effect mediated by baseline UPPS-P measures are included for all significant paths as numbers in circles (i.e., negative urgency accounted for 5.3% of the association between negative urgency PGS and nicotine use initiation).

**Supplementary Figure 4. Baseline UPPS-P mediating positive urgency PGS association with nicotine use initiation.**

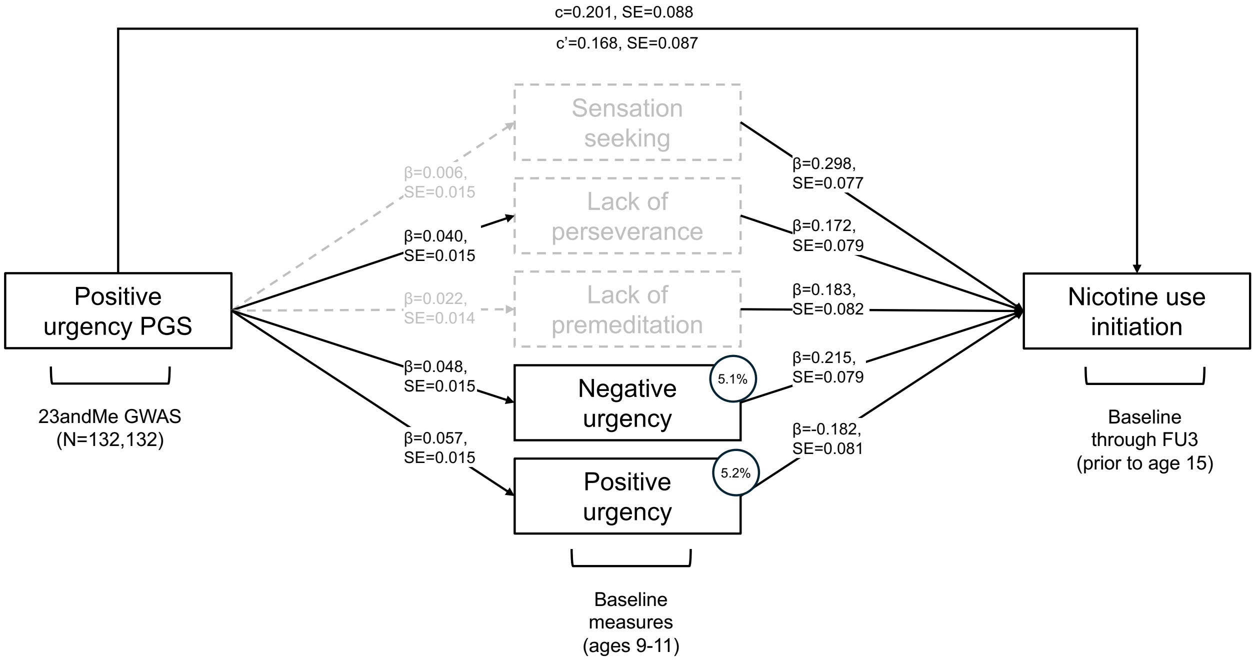


PGS = polygenic score; GWAS = genome-wide association study; c = total effect; c' = direct effect; FU = follow-up. Standardized regression coefficients (β) and standard errors (SE) are reported along each path. Grey dashed lines indicate individual paths and/or mediation effects with 95% CIs overlapping with 0 (i.e., non-significant). Estimates of proportion of the total effect mediated by baseline UPPS-P measures are included for all significant paths as numbers in circles (i.e., negative urgency and positive urgency accounted for 5.1 and 5.2% of association between positive urgency PGS and nicotine use initiation, respectively).
